# Health damages and disparities from municipal and medical waste incineration in Baltimore, USA

**DOI:** 10.1101/2025.06.27.25330313

**Authors:** Kevin J. Tu, Christopher D. Heaney, Greg Sawtell, Carlos Sanchez, Bonita Salmerón, Matthew A. Aubourg, Shiladitya DasSarma

**Author notes:** **Corresponding Author:** Kevin Tu.

## Abstract

**Background:** Waste incineration in Baltimore, USA, involves two major facilities: a municipal solid waste incinerator (WIN Waste) and the nation’s largest medical waste incinerator (Curtis Bay Medical Waste Incinerator). Both operate in socio-economically disadvantaged communities, raising concerns about cumulative environmental exposures and health disparities from hazardous air pollutants.

**Methods:** We estimated health impacts from available criteria incinerator emissions data (PM, NOx, SO_2_, CO). We used AERMOD to model ground-level pollutant concentrations, linked these to U.S. Census tracts, and monetized health damages using established relative risk and cost of illness data. Health disparities were evaluated by modeling incinerator-attributable mortality against the Social Vulnerability Index (SVI).

**Findings:** In 2024, the WIN Waste incinerator caused an estimated $53.8 million in health damages in Maryland and Washington DC. On average, the Curtis Bay Medical Waste Incinerator releases black smoke emissions for 52.5 minutes/day, a regulatory violation. The facility causes $36.9 million/year in health damages and is permitted to burn enough waste to cause up to $107.1 million/year; enforcing pollution controls at this site could prevent $13.8 million in annual harm in Baltimore. Combined, the two incinerators cause $97.0 million in damages annually. All-cause mortality from incinerator pollution was more common in communities with higher socioeconomic vulnerability.

**Interpretation:** Despite being required to install new pollution control equipment following community and regulatory pressure, the WIN Waste incinerator still causes significant health damages to Maryland and Washington DC. Meanwhile, repeated black smoke emissions from the Curtis Bay incinerator indicate that ongoing, uncontrolled pollution is also a major threat to public health in the region. Health damages from these incinerators disproportionately affect communities least able to bear the economic burden. Our conservative estimates highlight the need for urgent policy reforms, including stricter emissions monitoring, phasing out non-essential incineration, and ongoing cumulative impact assessments.

**Funding:** KJT and SD were funded by the University of Maryland Baltimore Provost’s Climate Health & Resilience internship program. KJT was supported by a Point Foundation Internship & Professional Development Award and the Alpha Omega Alpha Carolyn L. Kuckein Student Research Fellowship. BS, MAA, and CDH were supported by the National Institute of Environmental Health Sciences (NIEHS) P30 Center for Community Health: Addressing Regional Maryland Environmental Determinants of Disease (CHARMED) [grant no. P30ES032756]. BS, MAA, and CDH were supported by the Johns Hopkins Community Science and Innovation for Environmental Justice (CSI EJ) Initiative. CDH was supported by the National Institute for Occupational Safety and Health (NIOSH) Education and Research Center [grant no. T42OH0008428]. GS and CS are affiliated with the Curtis Bay Community Association and South Baltimore Community Land Trust. The authors have no other conflicts of interest to report.

## Introduction

Waste incineration releases a complex mixture of hazardous air pollutants that directly cause adverse health impacts, particularly through particulate matter (PM) and nitrogen oxides (NOx) (1). PM, especially fine particles (PM_2.5_), can penetrate deep into the lungs and bloodstream, triggering inflammation, exacerbating respiratory conditions such as asthma and chronic obstructive pulmonary disease (COPD), and increasing the risk of cardiovascular events including heart attacks and strokes (2). NOx contributes to the formation of ground-level ozone and secondary particulate matter, having been linked to asthma and further impairing lung function and aggravating respiratory diseases (3). Chronic exposure to these pollutants disproportionately affects vulnerable populations living near incinerators and other industrial sources (4). Here, we quantify the health damages stemming from two major incineration facilities operating in South Baltimore, Maryland: a large municipal solid waste (MSW) incinerator and the largest medical waste incinerator in the United States (U.S.).

The WIN Waste Innovations Baltimore facility (formerly Wheelabrator Baltimore), one of the U.S.’s largest MSW incinerators, is located in Westport, South Baltimore, a predominantly African-American community facing persistent socio-economic challenges including crime, housing abandonment, and unemployment (5). A previous report estimated that health damages attributed to the WIN Waste incinerator’s PM_2.5_ emissions alone amount to at least $55 million annually in Maryland and neighboring states (6). This estimate was based on the average emissions from 2014, 2015, and 2016 and is valued in 2010 U.S. dollars.

The Curtis Bay Medical Waste Incinerator, the largest of its kind in the U.S., is situated in the eponymous South Baltimore neighborhood, a low-income, heavily industrialized corridor along the Patapsco River (7). The incinerator has a documented history of regulatory non-compliance, including a $1.75 million state penalty in 2023 for multiple permit violations and ongoing legal challenges (8). Such operational deficiencies, coupled with the hazardous nature of medical waste, may potentiate local health risks and contribute to cumulative environmental exposures in an already overburdened area.

Situated within communities already burdened with industrial pollution, the proximity of these two incinerators and their emissions raises profound concerns regarding cumulative environmental exposures and their impact on health disparities (9). We partnered with the South Baltimore Community Land Trust, a community nonprofit in South Baltimore, to quantify incinerator-attributable mortality and morbidity of cardiovascular, respiratory, and metabolic disease via atmospheric and health impact modeling. Our findings may inform evidence-based policy, empower local organizations, and improve healthcare providers’ understanding of how medical waste incineration directly impacts patient and community health. More broadly, this research underscores the need for cleaner waste management and monitoring to safeguard vulnerable populations and address health inequities.

## Methods

### Emissions Estimates and Smokestack Constants

Annual emissions from the WIN Waste Baltimore incinerator were obtained via Maryland Public Information Act requests to the Maryland Department of the Environment. For the Curtis Bay Medical Waste Incinerator, because reported emissions data are scarce, emissions were estimated for two scenarios: typical throughput (26,000 tons/year) and maximum permitted throughput (56,000 tons/year) (10). We focused on PM_2.5_, NOx, SO_2_, and CO because they were among the few high-volume criteria pollutants reported by the incinerators and available through EPA emissions data. Emissions were calculated by multiplying throughput by U.S. EPA AP-42 emission factors for medical waste incinerators, including criteria pollutants, metals, acid gases, and organics (11). Smokestack height and diameter, plume temperature, and emission velocity parameters for the WIN Waste Baltimore incinerator were obtained via previously published reports (12). As the same for the Curtis Bay Medical Waste Incinerator is not readily available within the public record, they were obtained via minimum building code requirements for medical waste incinerators or taken from standard estimates from similar incinerators (13). The constants used in this study are available in **Table S1**. Where AP-42 emission factors were unavailable, we applied control efficiency estimates based on peer-reviewed retrofit studies and regulatory guidance, assuming a 50% reduction for NOx (14).

### Estimating Ground-Level Pollutant Concentrations

Ground-level concentrations (µg/m^3^) of pollutants were predicted using the United States Environmental Protection Agency’s (U.S. EPA) AERMOD modeling system (version 24142) (15). AERMOD is a steady-state Gaussian plume model widely recognized for regulatory air dispersion modeling, estimating pollutant concentrations from various sources while accounting for meteorological conditions, terrain, and building downwash effects.

Hourly meteorological data for AERMOD were prepared using the AERMET (AERMOD Meteorological Preprocessor) component of the modeling system. Surface meteorological data were obtained from the nearest airport, the Baltimore/Washington International Thurgood Marshall Airport. Upper air meteorological data were derived from the nearest upper air monitoring station, the Dulles International Airport/Sterling Upper Air Meteorological station. These raw meteorological data undergo processing within AERMET to generate the boundary layer parameters required by AERMOD. Surface characteristics were determined using the AERSURFACE pre-processor. AERSURFACE utilizes land cover data from the U.S. Geological Survey National Land Cover Database. For both meteorological and land cover data, we used information from the same year as the emissions and modeling whenever available. However, because land cover data was only available through 2023, we used the 2023 land cover data for modeling emissions from 2024.

Terrain elevation data were incorporated into the model using AERMAP (AERMOD Terrain Preprocessor). Elevation data were obtained from NASA’s Shuttle Radar Topography Mission USA 30M dataset. AERMAP processes this data to provide terrain height and receptor elevations for use in AERMOD. Elevation data were processed through AERMAP to ensure proper receptor height determination. The influence of nearby structures, specifically the incinerator building, on pollutant dispersion was assessed using the BPIP (Building Profile Input Program) pre-processor. Dimensional data for the incinerator building were determined through the OpenStreetMap database. BPIP calculates building dimensions and orientation to account for building downwash effects, which may impact pollutant concentrations in the immediate vicinity of structures.

AERMOD was run using regulatory default options. A 50 km radius was established as the modeling domain, representing the outer limit of AERMOD’s typical applicability for predicting concentrations. An urban population of 565,000, simulating the population of Baltimore, was used to define the urban dispersion coefficient. A flagpole height of 2 meters was used for receptor height, representing typical breathing zone height. Annual average pollutant concentrations were determined for the grid area by taking the maximum calculated value.

Separate AERMOD simulations were conducted for each year of available WIN Waste data. For the Curtis Bay Medical Waste Incinerator, separate models were run for each condition (i.e., typical controlled, typical uncontrolled, maximum controlled, maximum uncontrolled).

Additionally, a combined model for both incinerators operating in 2024 was run, utilizing the typical operating conditions for the Curtis Bay facility.

### Population Exposure Assessment

To quantify population exposure, ground-level pollutant concentrations estimated by AERMOD were spatially joined with U.S. Census Bureau tract boundaries for Maryland and Washington DC. For census tracts containing one or more AERMOD grid points, the average pollutant concentration within that tract was assigned as the representative exposure value. For tracts without direct grid points, the concentration from the nearest AERMOD grid point was assigned to ensure comprehensive coverage. While pollution dispersion likely extended to other states as previously described, this analysis was limited to Maryland and Washington DC due to technical limitations inherent in AERMOD’s modeling capabilities (6).

### Health Impact Assessment

The health endpoints quantified in this study were: all-cause mortality, asthma cases, diabetes, ischemic heart disease, stroke, and COPD. These conditions were selected due to their well-characterized epidemiological links to air pollution exposure, though it is important to note that other potential health impacts not included in this list may also be associated with incinerator emissions. Health impacts from exposure to incinerator emissions were estimated using health endpoint-specific relative risks (RR) derived from U.S. EPA assessments or large meta-analyses, available in **Table S2** (16–27). These relative risks (RR) were converted to β coefficients using **Equation 1**

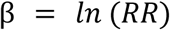

**Equation 1**. Conversion of RR to β coefficients

For each census tract, the number of additional health cases for a given endpoint was calculated using **Equation 2** (28):

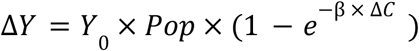

**Equation 2**. Health impact function to determine additional cases of a particular endpoint

Where ΔY refers to the number of additional cases, Y_0_ refers to the incidence or prevalence of a specific health endpoint per 100,000 population, Pop refers to the population, and ΔC refers to the increase in concentration (μg/m^3^) of a pollutant as a result of the incinerator estimated for each census tract. Baseline rates were sourced from the National Institutes of Health, the Centers for Disease Control and Prevention, and the Global Burden of Disease study, corresponding to the year of analysis (**Table S3**). Population was obtained from the U.S. Census Bureau’s

American Community Survey (ACS) 5-year estimates. For the WIN Waste analysis, American Community Survey (ACS) 5-year estimates corresponding to the year of analysis were utilized. For instances where 2024 data was required and unavailable, 2023 ACS data was used as a proxy.“Concentration” represents the ground-level pollutant concentration in mass per unit volume.

### Economic Valuation

Economic costs associated with health impacts were calculated by multiplying the estimated additional cases for each health endpoint by their respective per-case economic valuations. Mortality benefits were monetized using the EPA’s 2024 Value of a Statistical Life ($11.74 million in 2025 dollars) (29), and morbidity benefits were monetized using previously reported values (**Table S4**). When possible, inflation was normalized to and reported in 2025 dollar values using the Bureau of Labor Statistics’ Consumer Price Index.

### Disparities Assessment

To assess disparities in health impacts in 2024 data, we integrated estimated health outcomes with the Social Vulnerability Index (SVI). The SVI, a measure developed by the U.S. Centers for Disease Control and Prevention (CDC), quantifies social vulnerability based on 16 census variables grouped into four themes: Socioeconomic Status, Household Characteristics, Racial and Ethnic Minority Status, and Housing Type and Transportation (30). A higher SVI indicates greater social vulnerability. Most recent SVI data from 2022 were obtained from the CDC/ATSDR database and joined with health impact data at the census tract level using the unique FIPS code. All-cause mortality, being the most significant cause of health damages across all tested scenarios, was selected as the primary outcome variable for these analyses. We conducted univariate generalized linear models (GLMs) with a Gaussian family for each of the 16 SVI census variables against the rate of all-cause mortality, stratified by incineration condition. Subsequently, significant SVI variables (p < 0.05) from the univariate analyses were included in multivariate GLMs, which also adjusted for pollutant concentration and total population.

### Software

The AERMOD workflow was conducted using AERMET View (version 13.0) and AERMOD View (version 13.0). Population, health, and economic damage modeling was accomplished using R (version 4.4.1). A p-value of <0.05 was considered significant.

## Results

### Health Damages from WIN Waste Municipal Waste Incinerator

Analysis of air pollutants including PM, NOx, sulfur dioxide (SO_2_), and carbon monoxide (CO) from the WIN Waste Baltimore Incinerator revealed significant health damages. Air pollution dispersion, characterized using AERMOD, extended widely across the study area, with the highest concentrations centered east of the incinerator near the Midtown and Southeast regions of Baltimore (**Figure 1A-B**). This dispersion resulted in quantifiable health damages across seven endpoints: all-cause mortality, ischemic heart disease, asthma, respiratory tract cancer, stroke, COPD, and diabetes.

**Figure 1.**
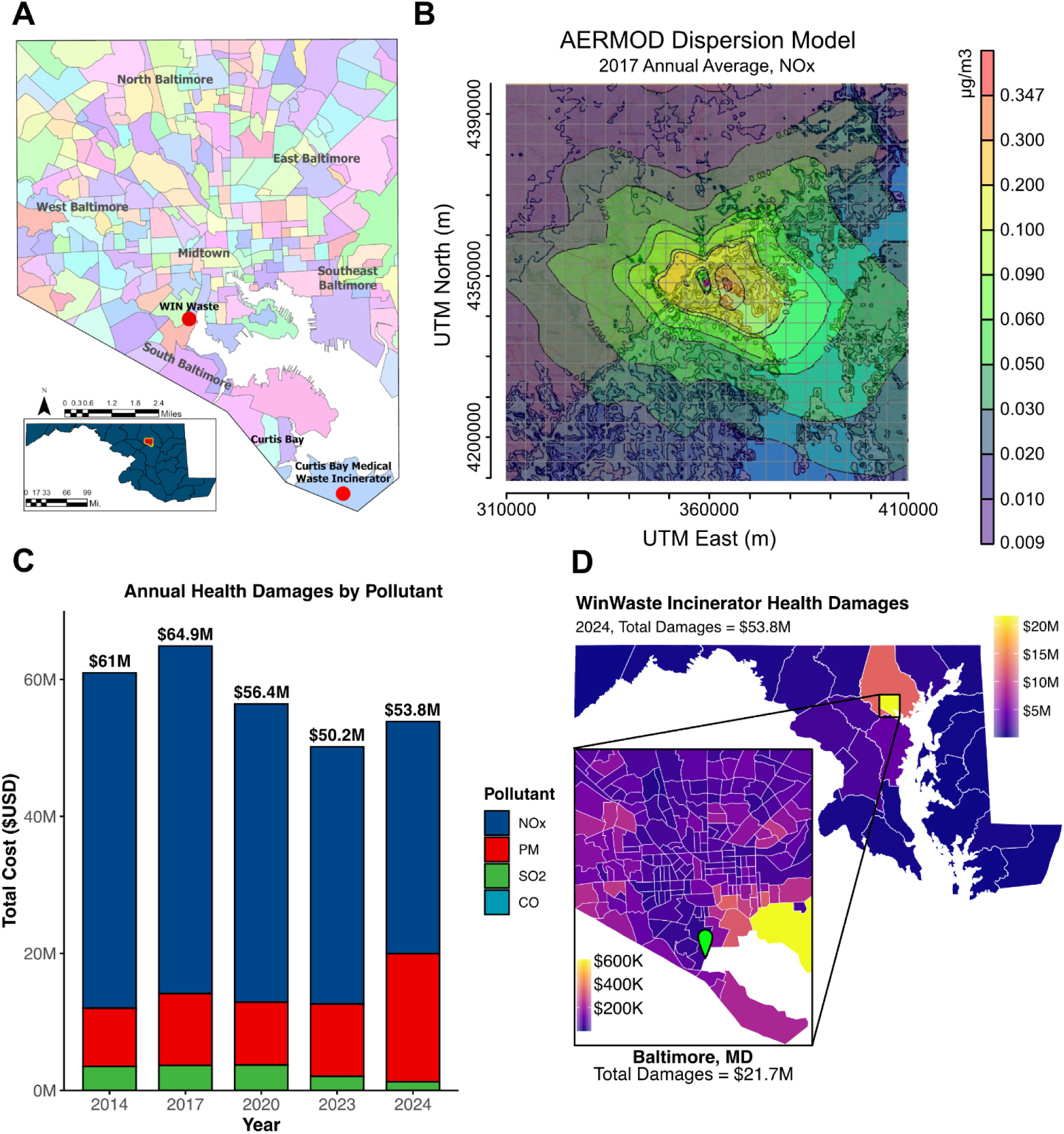
Monetized health damages from WIN Waste Baltimore Incinerator (2014-2024). A) Map of Baltimore City, MD U.S. Neighborhood Statistical Areas and Geographic Areas, in relation to the WIN Waste and Curtis Bay Medical Waste Incinerator facilities, indicated by red points. B) Representative image of annualized average air pollution dispersion from the WIN Waste Baltimore incinerator, scale indicates increased pollution concentration as a result of the incinerator in µg/m^3^ (NOx 2017). C) Stacked bar chart of annual health-related economic damages categorized by pollutant 2014-2024; the damages from CO are not visually distinguishable in the chart due to being relatively low, but they are accounted for in the annual totals. D) Health economic damages ($USD) by Maryland/Washington DC county and Baltimore census tracts in 2024. Green pin represents the location of the WIN Waste incinerator. PM = particulate matter, NOₓ = Nitrogen Oxides, SO_2_ = Sulfur Dioxide, CO = Carbon Monoxide.

NOx emissions were identified as the primary driver of mortality cases attributable to the WIN Waste incinerator, followed in impact by PM, SO₂, and CO, respectively (**Table 1**). Peak health damages reached $64.9 million in 2017, subsequently decreasing to $53.8 million in 2024, with a low of $50.2 million in 2023 (**Figure 1C**). This reduction is likely linked to the onset of community action in 2017 and subsequent local policy changes. While NOx emissions and associated mortality decreased in this time, PM emissions and their related damages have risen since 2020, with a notable increase in 2024 (**Figure 1C**).

**Table 1.**
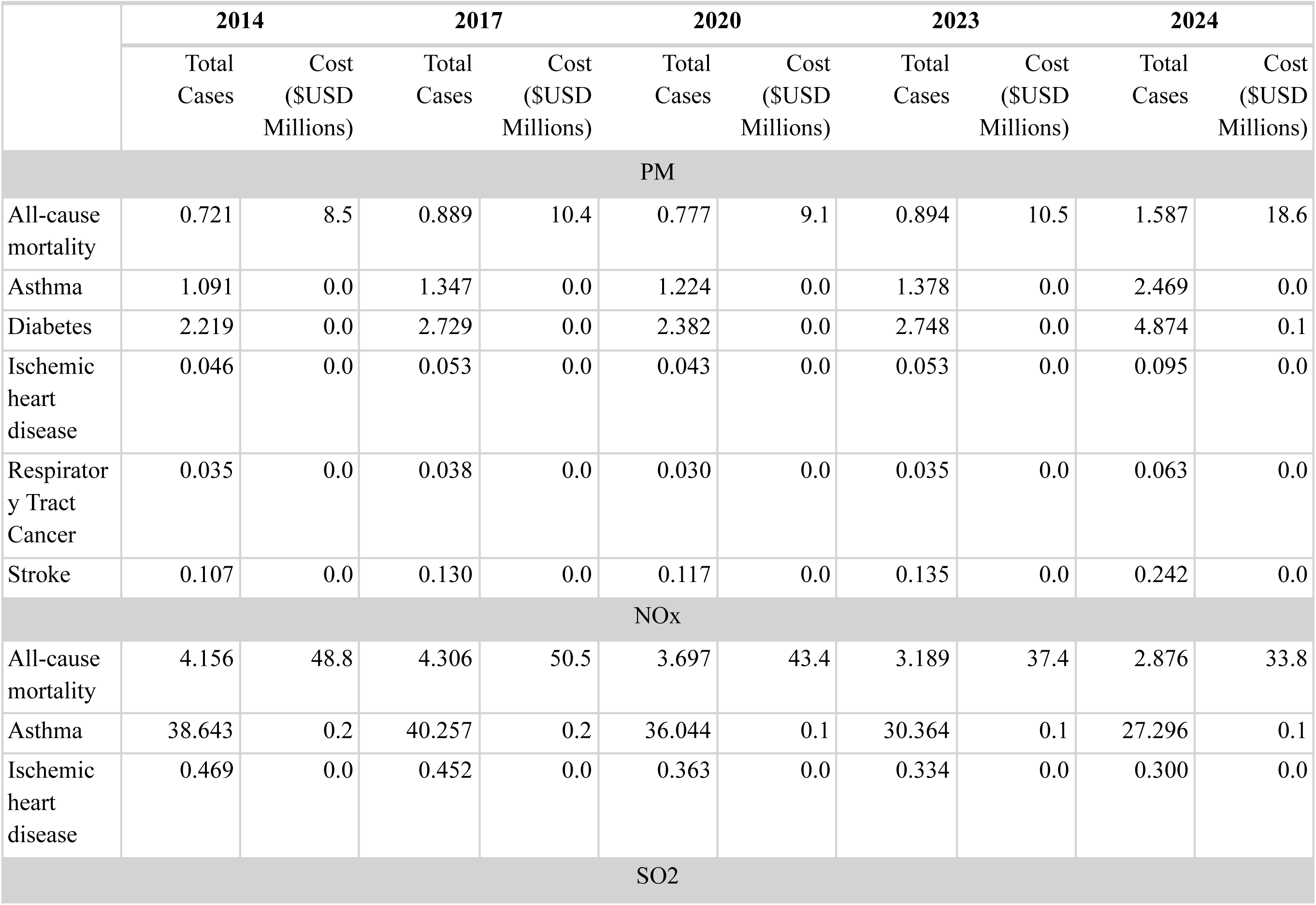
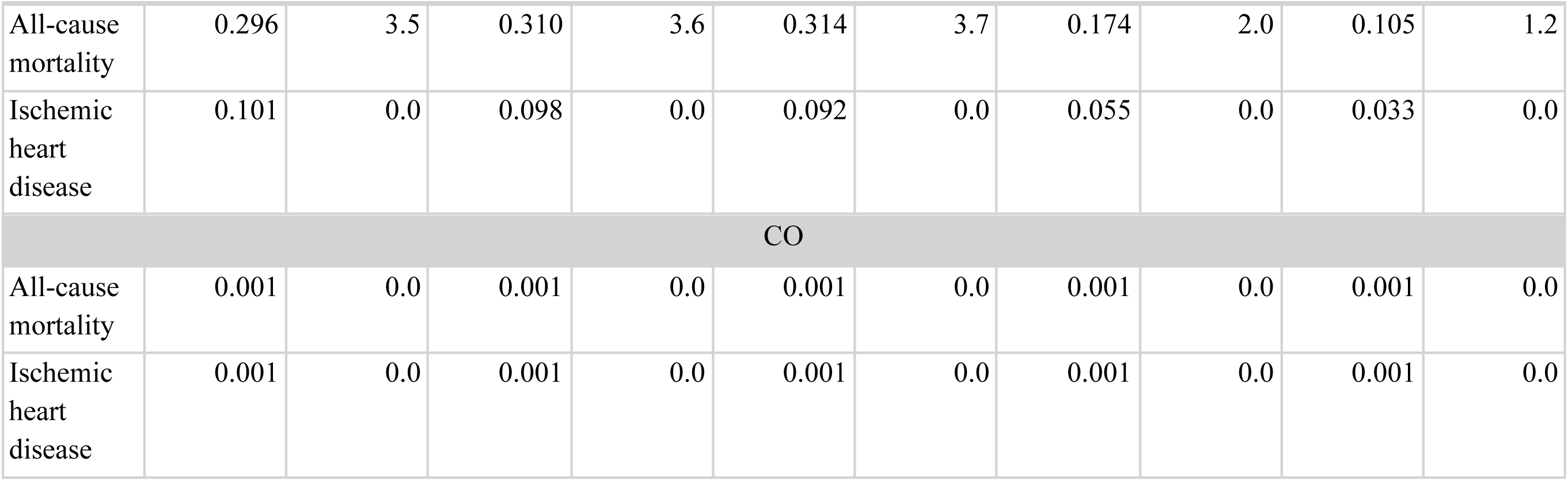
Annual Human Health Effects and Monetary Valuations Associated With emissions from the WIN Waste Incinerator. PM: Particulate matter, NOₓ: Nitrogen Oxides, SO2: Sulfur Dioxide, CO: Carbon Monoxide, DSI: Dry Sorbent Injection, CI: Activated Carbon Injection, FF: Fabric Filter

The majority of damages were concentrated in Baltimore City ($21.7 million in 2024) and surrounding Baltimore County, though impacts extended to Washington DC and potentially other states (**Figure 1D**). Within Baltimore City, the most affected areas were southeastern census tracts and West Baltimore, consistent with the location of peak NOx concentration from our AERMOD analysis.

### Health Damages from the Curtis Bay Medical Waste Incinerator

The Curtis Bay Medical Waste Incinerator has a longstanding record of environmental crimes, including improper waste burning, inadequate maintenance of pollution control systems, and illegal discharges into local waterways (8). Our analysis estimates health damages from air emissions under two scenarios: (1) typical annual throughput of 26,000 tons and (2) the facility’s maximum permitted throughput of 56,000 tons.

Trail camera footage collected by the South Baltimore Community Land Trust shows black smoke emissions averaging 52.5 minutes/day, even after the State of Maryland filed a lawsuit against the incinerator for air quality violations (**Video 1**) (31). These findings suggest continued noncompliance with emissions passing through air vents instead of pollution control equipment. To reflect this, emissions were modeled under two bounds: a lower-bound scenario assuming all reported pollution controls—dry sorbent injection (DSI), activated carbon injection (ACI), and fabric filtration (FF)—are fully operational (“controlled”), and an upper-bound scenario assuming no control technologies are functioning (“uncontrolled”). If actual emissions fall within this range, they may more likely fall closer to the upper bound, given the facility’s history of black smoke events. Emission estimates for both scenarios were generated by multiplying annual throughput by U.S. EPA AP-42 emission factors for criteria pollutants.

Of note, this analysis cannot precisely model the observed black smoke itself due to a lack of quantifiable emissions rates. This smoke is composed of black and/or brown carbon, both of which have substantial health impacts; thus, our estimates likely do not encapsulate its whole impact (32).

Dispersion modeling for the Curtis Bay incinerator showed that pollutant concentrations were highly localized, with the highest levels concentrated in the immediate vicinity (∼1 km) of the facility, particularly in the Curtis Bay neighborhood (**Figure 2A**). As a result, fewer people were exposed overall compared to WIN Waste, but those who were exposed experienced much higher pollutant concentrations.

**Figure 2.**
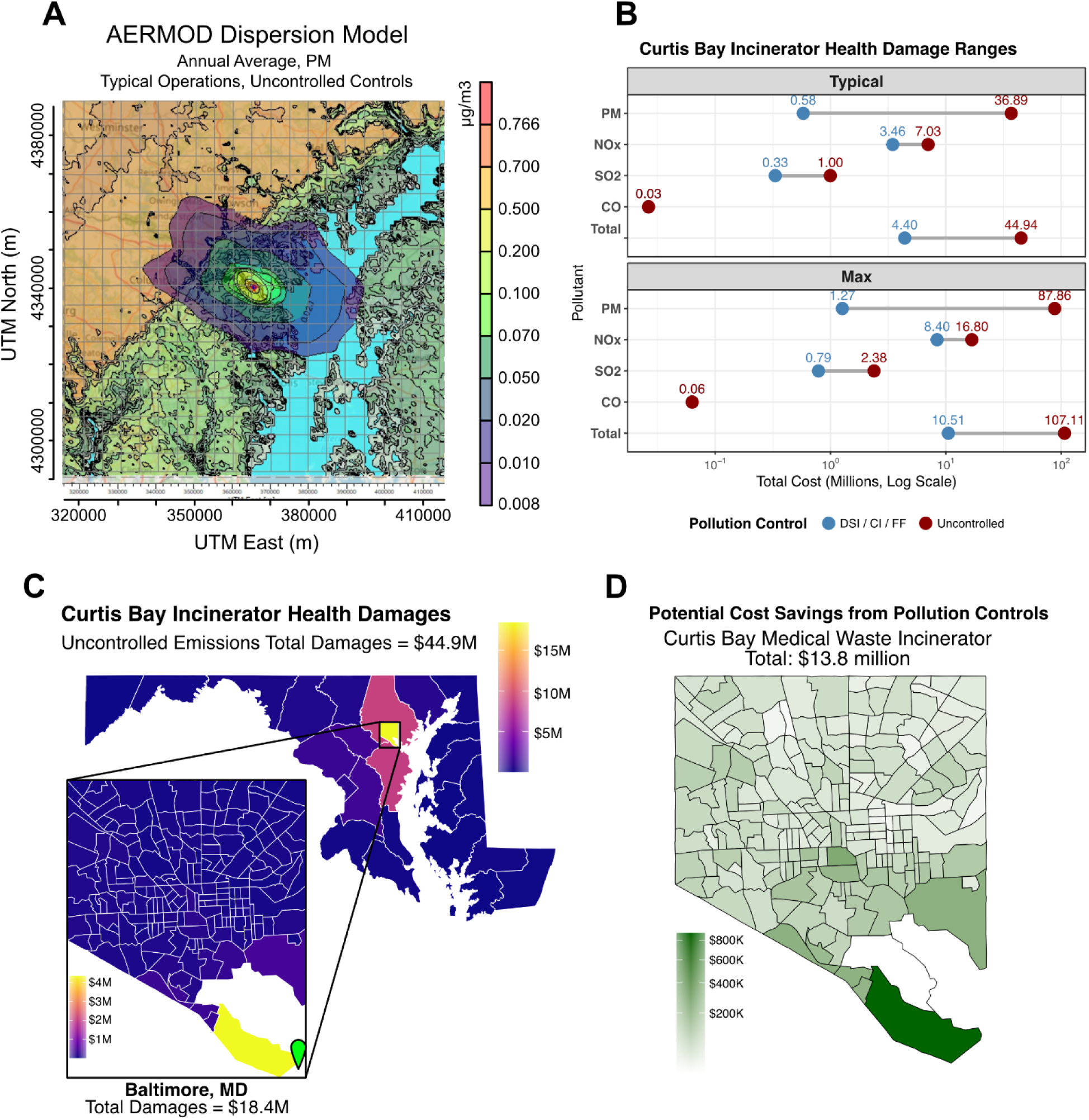
Health damages and potential cost savings from the Curtis Bay Medical Waste Incinerator. A) Representative image of annualized average air pollution dispersion from the Curtis Bay Medical Waste Incinerator, scale indicates increased pollution concentration as a result of the incinerator in µg/m^3^ (PM, typical operations, uncontrolled controls). B) Health damage ranges by pollutant under typical and maximum throughput scenarios, categorized by pollution control (controlled vs. uncontrolled). C) Uncontrolled emissions total health damages by Maryland/Washington DC county and Baltimore census tracts. D) Potential cost savings from implementing pollution controls at the Curtis Bay Medical Waste Incinerator, by Baltimore census tracts. DSI: Dry Sorbent Injection, CI: Activated Carbon Injection, FF: Fabric Filter

**Video 1.**
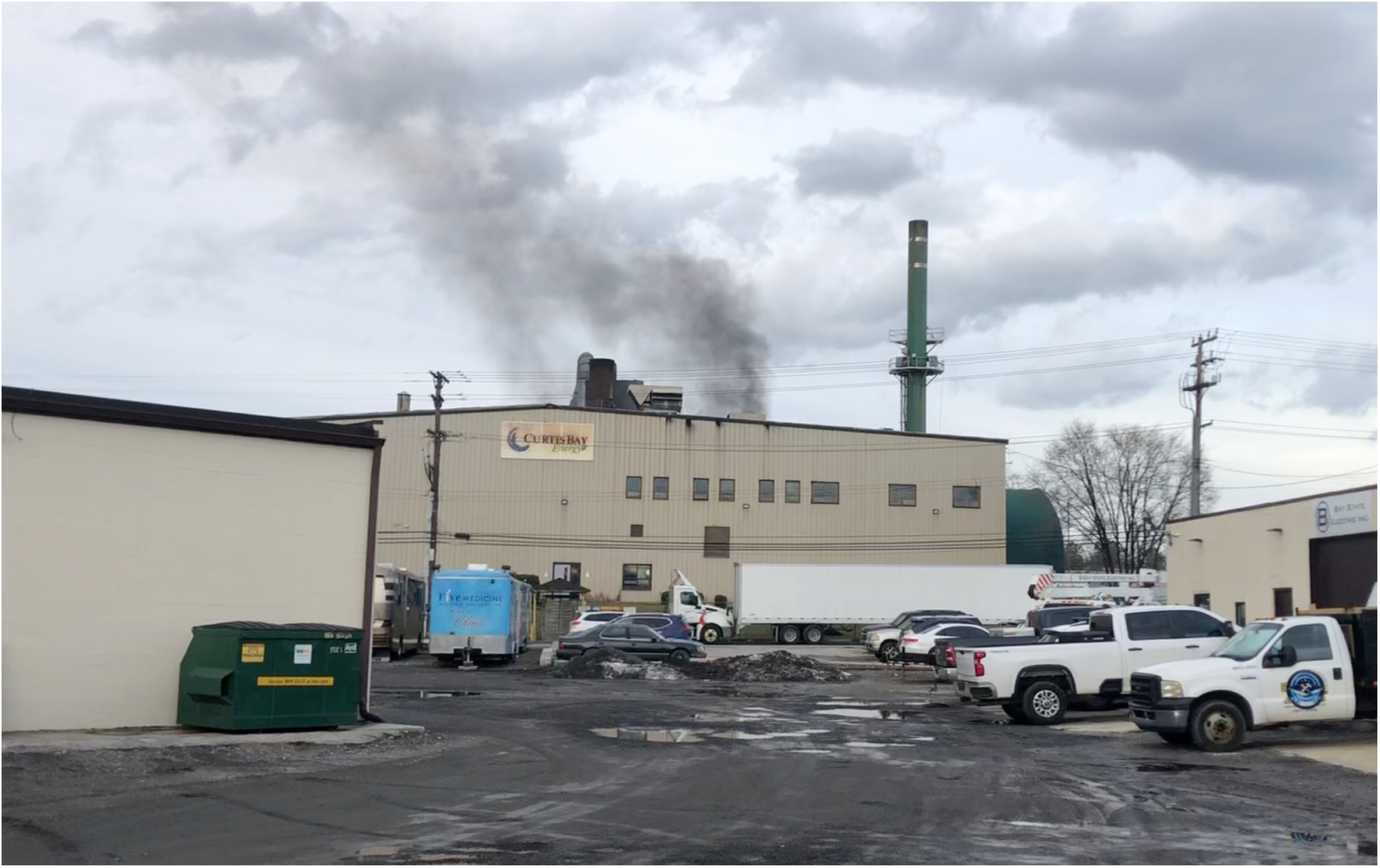
Example footage of a black smoke event at the Curtis Bay Medical Waste Incinerator. Dated January 26, 2024; Source: South Baltimore Community Land Trust.

Under typical operating conditions, health damages from the Curtis Bay incinerator ranged from $0.58 million to $36.9 million, depending on the level of operational pollution controls. However, at maximum permitted capacity, damages could escalate to $10.51 million to $107.11 million (**Figure 2B**). Given the facility’s history of black smoke events, the true damages may be closer to the upper bound of these estimates. These damages are predominantly borne by Baltimore City, with additional impacts on Baltimore County and Anne Arundel County. Within Baltimore City, the Curtis Bay neighborhood experienced the vast majority of these damages (**Figure 2C**). PM emissions were the primary driver of damages in the uncontrolled scenario, while NOx was the primary driver in the controlled scenario (**Table 2**).

**Table 2.** Health endpoint cases and economic damages from the Curtis Bay Medical Waste Incinerator. PM: Particulate matter, NOₓ: Nitrogen Oxides, SO2: Sulfur Dioxide, CO: Carbon Monoxide, DSI: Dry Sorbent Injection, CI: Activated Carbon Injection, FF: Fabric Filter

|  | Typical Controlled<br>(DSI/CI/FF) |  | Typical Uncontrolled |  | Max Controlled<br>(DSI/CI/FF) |  | Max Uncontrolled |  |
| --- | --- | --- | --- | --- | --- | --- | --- | --- |
| | Total Cases | Cost (\$USD Millions) | Total Cases | Cost (\$USD Millions) | Total Cases | Cost (\$USD Millions) | Total Cases | Cost (\$USD Millions) |
| PM |  |  |  |  |  |  |  |  |
| All-cause mortality | 0.049 | 0.6 | 3.128 | 36.7 | 0.107 | 1.3 | 7.450 | 87.5 |
| Asthma | 0.076 | 0.0 | 4.839 | 0.0 | 0.166 | 0.0 | 11.525 | 0.0 |
| Diabetes | 0.150 | 0.0 | 9.561 | 0.1 | 0.329 | 0.0 | 22.744 | 0.3 |
| Ischemic heart disease | 0.003 | 0.0 | 0.187 | 0.0 | 0.006 | 0.0 | 0.445 | 0.0 |
| Respiratory Tract Cancer | 0.002 | 0.0 | 0.123 | 0.0 | 0.004 | 0.0 | 0.293 | 0.0 |
| Stroke | 0.007 | 0.0 | 0.476 | 0.0 | 0.016 | 0.0 | 1.133 | 0.1 |
| NO<sub>x</sub> |  |  |  |  |  |  |  |  |
| All-cause mortality | 0.294 | 3.4 | 0.596 | 7.0 | 0.713 | 8.4 | 1.426 | 16.7 |
| Asthma | 2.750 | 0.0 | 5.687 | 0.0 | 6.798 | 0.0 | 13.590 | 0.1 |
| Ischemic heart disease | 0.030 | 0.0 | 0.063 | 0.0 | 0.075 | 0.0 | 0.149 | 0.0 |
| SO<sub>2</sub> |  |  |  |  |  |  |  |  |
| All-cause mortality | 0.028 | 0.3 | 0.085 | 1.0 | 0.067 | 0.8 | 0.203 | 2.4 |
| Ischemic heart disease | 0.009 | 0.0 | 0.027 | 0.0 | 0.021 | 0.0 | 0.064 | 0.0 |
| CO |  |  |  |  |  |  |  |  |
| All-cause mortality | 0.002 | 0.0 | 0.002 | 0.0 | 0.005 | 0.1 | 0.005 | 0.1 |
| Ischemic heart disease | 0.002 | 0.0 | 0.002 | 0.0 | 0.005 | 0.0 | 0.005 | 0.0 |

Modeling the impact of implementing working controls, as required by regulations, revealed a potential prevention of $13.8 million in health damages for Baltimore alone, with over $800,000 in potential savings specifically for the Curtis Bay neighborhood (**Figure 2D**). This highlights the economic rationale for enforcing stricter operational standards.

### Combined Damages and Disparities of Harm

Using AERMOD, we modeled air pollution dispersion using the two incinerators together, and we find that, with uncontrolled emissions from Curtis Bay, the total health damages to Maryland/Washington DC alone reached $97.0 million (**Table 3**). These damages are largely localized to Baltimore, but also significantly affect Baltimore County, Anne Arundel County, central Maryland counties (Howard County, Prince George’s County, Montgomery County), and Washington DC (**Figure 3A**). Baltimore bore the greatest proportion of harm, particularly West, South, and Southeast Baltimore (**Figure 3A**).

**Figure 3.**
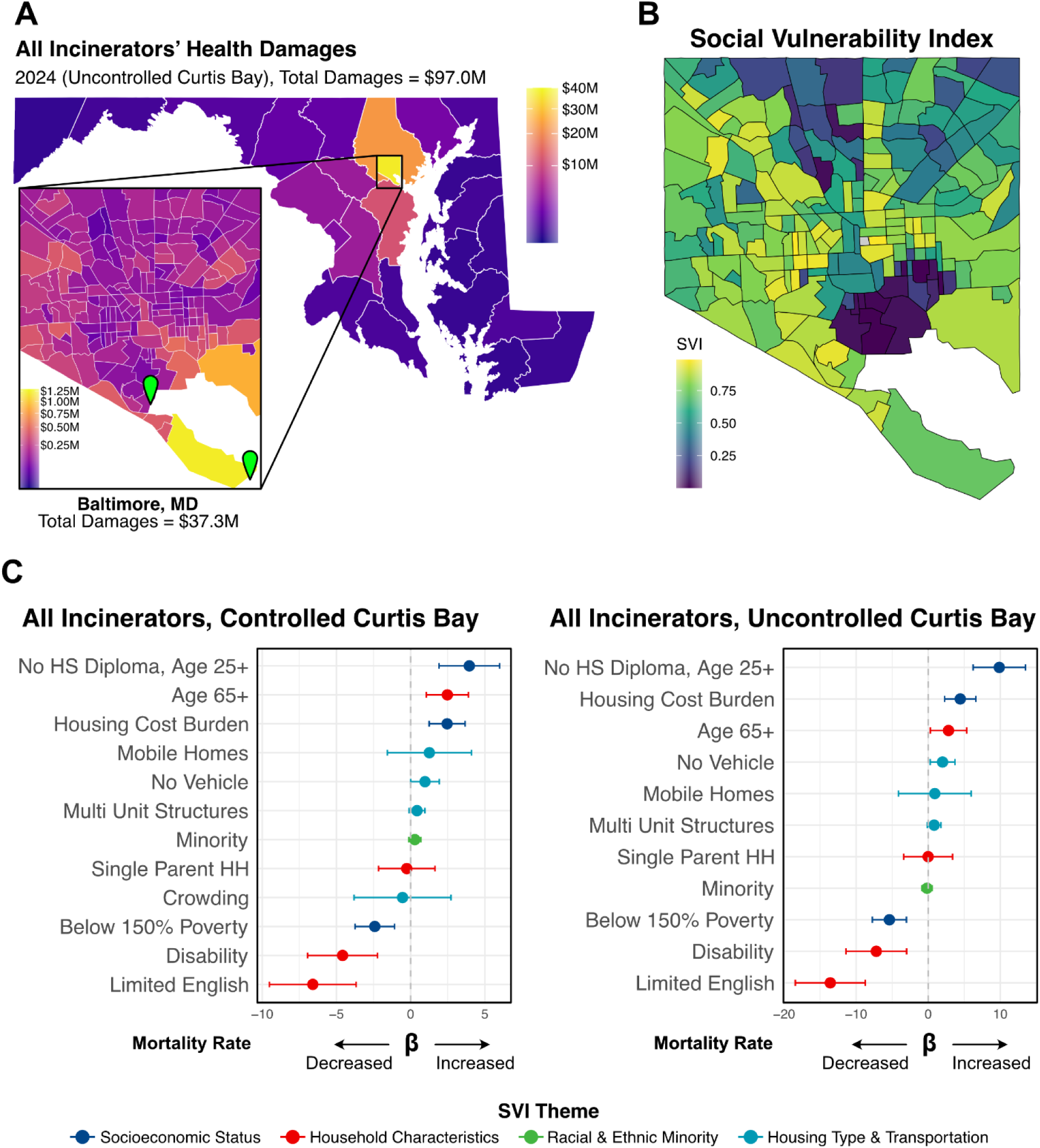
Cumulative health damages, their spatial distribution in Baltimore, and associations with social vulnerability. A) Map of total cumulative health damages (in USD) from all incinerators (2024, uncontrolled Curtis Bay), with a focus on breakdown in Baltimore City. B) Social Vulnerability Index (SVI) map of Baltimore. C) Regression coefficients (β) and 95% confidence intervals for the association between social vulnerability index (SVI) variables and all-cause mortality due to incinerator pollution, for both controlled and uncontrolled Curtis Bay scenarios.

**Table 3.** Annual Human Health Effects and Monetary Valuations Associated With emissions from both the WIN Waste and Curtis Bay Medical Waste Incinerators. PM: Particulate matter, NOₓ: Nitrogen Oxides, SO2: Sulfur Dioxide, CO: Carbon Monoxide, DSI: Dry Sorbent Injection, CI: Activated Carbon Injection, FF: Fabric Filter

|  | Typical Controlled<br>(DSI/CI/FF) |  | Typical Uncontrolled |  |
| --- | --- | --- | --- | --- |
| | Total Cases | Cost (\$USD Millions) | Total Cases | Cost (\$USD Millions) |
| PM |  |  |  |  |
| All-cause mortality | 1.752 | 20.6 | 4.518 | 53.0 |
| Asthma | 2.722 | 0.0 | 7.050 | 0.0 |
| Diabetes | 5.378 | 0.1 | 13.854 | 0.2 |
| Ischemic heart disease | 0.105 | 0.0 | 0.272 | 0.0 |
| Respiratory Tract Cancer | 0.069 | 0.0 | 0.179 | 0.0 |
| Stroke | 0.267 | 0.0 | 0.693 | 0.0 |
| NOx |  |  |  |  |
| All-cause mortality | 3.216 | 37.8 | 3.482 | 40.9 |
| Asthma | 30.820 | 0.1 | 33.386 | 0.1 |
| Ischemic heart disease | 0.339 | 0.0 | 0.367 | 0.0 |
| SO2 |  |  |  |  |
| All-cause mortality | 0.160 | 1.9 | 0.224 | 2.6 |
| Ischemic heart disease | 0.052 | 0.0 | 0.073 | 0.0 |
| CO |  |  |  |  |
| All-cause mortality | 0.003 | 0.0 | 0.003 | 0.0 |
| Ischemic heart disease | 0.003 | 0.0 | 0.003 | 0.0 |

The Social Vulnerability Index (SVI), a US CDC measure for quantifying vulnerability across 16 census variables in four themes (socioeconomic status, household characteristics, racial and ethnic minority, housing type & transportation), showed a spatial correlation with the distribution of health damages in Baltimore. Namely, areas with higher SVI, such as West Baltimore and South Baltimore, experienced greater damage (**Figure 3B**). This aligns with environmental justice literature suggesting that vulnerable populations are disproportionately affected by environmental pollution due to co-location near industrial facilities, a phenomenon often described in Baltimore as “the black butterfly” (33).

Univariate regressions of the 16 SVI census variables as percentages with all-cause mortality rate per 100,000, chosen as a proxy for overall health damages, were conducted for both controlled and uncontrolled Curtis Bay scenarios (**Table S5**). The significant variables were then used within a multivariable regression. Socioeconomic vulnerabilities, such as a lack of a high school diploma (Uncontrolled: β 9.86, 95% CI 6.23 - 13.49; Controlled: β 3.95, 95% CI 1.91 - 6.00) and high housing cost burden (Uncontrolled: β 4.44, 95% CI 2.29 - 6.61; Controlled: β 2.46, 95% CI 1.24 - 3.67), were associated with higher mortality rates (**Figure 3C**). Conversely, household characteristics like limited English proficiency (Uncontrolled: β-13.58, 95% CI-18.44--8.725; Controlled: β-6.59, 95% CI-9.51--3.67) and disability (Uncontrolled: β-7.20, 95% CI-1.14--3.00; Controlled: β-4.59, 95% CI-6.94--2.23) were associated with less mortality (**Figure 3C**). This suggests that socioeconomically vulnerable populations bear a disproportionate burden of health damages from incinerator pollution, despite likely having the least capacity to afford such impacts. Interestingly, high levels of poverty were associated with less mortality from air pollution, indicating some heterogeneity in these findings (**Figure 3C**). The variables significant in both controlled and uncontrolled models were similar, but the disparities in vulnerability were widened by increased pollution in the uncontrolled model, suggesting that air pollution exacerbates existing disparities (**Figure 3C**). Furthermore, older adults (age 65+) were found to have significantly greater mortality, which aligns with expected epidemiological patterns and lends validity to the model.

## Discussion

We estimate annual health damages of approximately $54 million from WIN Waste Baltimore and $37 million from Curtis Bay, totaling $97 million in combined impacts. These figures are likely conservative for several reasons. First, these impacts were assessed only within Maryland, excluding neighboring states; previous analyses of the WIN Waste incinerator suggest that when considering the surrounding states, the actual costs may be 2.5-fold higher (6). Second, our analysis was limited to four major criteria pollutants (PM_2.5_, NO_x_, SO_2_, and CO) based on available emissions data. In reality, these incinerators also emit other harmful pollutants including heavy metals like lead, black and brown carbon, and volatile organic compounds that are strongly linked to additional health impacts but were not captured in our estimates (34,35).

Third, this study did not consider the downstream health effects of climate change as a result of the greenhouse gases released by the incinerator. And lastly, prior research indicates that incineration-related harm also results from pollutant ingestion through contaminated soil, water, or food pathways, which we were unable to model in this study (36).

Notably, we find that incinerator-related health damages are borne predominantly by socioeconomically vulnerable communities, who are the least equipped to handle these health damages and often have experienced the health impacts of industrial pollution for multiple generations. Historically, polluting infrastructure has disproportionately harmed vulnerable communities like South and West Baltimore (4). Moreover, formal health impact assessments for these operating facilities are rare, and a persistent lack of emissions monitoring and public data is a continuing problem. The Curtis Bay incinerator, for instance, operated for years with minimal transparency despite repeated violations. Our analysis indicates that simply enforcing baseline pollution controls at Curtis Bay to meet existing regulatory standards could prevent an estimated $13.8 million in annual health damages in Baltimore alone. However, our community trail camera data suggest such controls have not been implemented, despite ongoing lawsuits. The continued operation of the Curtis Bay incinerator under inadequate controls perpetuates environmental injustice and underscores the urgent need for community-centered reform.

Amid these systemic challenges, we also uncover a story of resilience: community action at WIN Waste Baltimore was associated with an estimated $11.1 million annualized health damage reduction by 2024, versus 2017 levels. This outcome is one of hard-won progress and community advocacy catalyzed by science. However, despite this reduction, the WIN Waste Baltimore continues to be the single largest source of harmful emissions in the city. Federal oversight, specifically the U.S. EPA’s standards for large MSW incinerators, has not been substantively updated since 2006, a delay of over a decade beyond the Clean Air Act’s mandated review cycle (37). This regulatory lag means that even with some local improvements, incinerators like WIN Waste Baltimore operate under standards that do not reflect the best available control technologies or current understandings of health impacts.

Our findings clearly suggest the need for policy action to protect human health in the region. First, Maryland and Baltimore City should mandate continuous emissions monitoring and public reporting for all incinerators. Second, policies should disincentivize medical waste incineration and promote safer alternatives, aligning with World Health Organization calls against non-essential incineration (38). Finally, state agencies should conduct rigorous cumulative impact assessments, including socioeconomic vulnerability and co-exposures, before permit renewals for high-emitting facilities. Importantly, while these are lessons drawn from a local context, they offer broadly applicable guidance for global waste and environmental health policy—particularly in communities facing similar patterns of pollution and structural vulnerability.

Furthermore, waste-generating institutions, especially healthcare facilities like hospitals, share responsibility for these health impacts and must adopt upstream waste reduction strategies through improved waste segregation and procurement reform. Incineration, particularly at Curtis Bay, should not remain the primary disposal method. If demand for waste disposal increases and the facility operates at its maximum permitted throughput, annual health damages could exceed $107.11 million. This public health threat is preventable; a local community nonprofit suggested Baltimore has ample alternative capacity (over 20 times current incineration volume) for safer treatment (39). Major waste-generating institutions committed to community health alongside policymakers should lead this transition, requiring vendor accountability, transparent audits, and investment in non-burn alternatives.

Future research should integrate real-time air monitoring, longitudinal health data, and cost-benefit analyses of alternatives. However, current evidence is sufficient for immediate action; consequently, policymakers and decision-makers should re-evaluate and eliminate incineration as the default status quo method of disposal.

## Data availability

All data and code used in this study are available at https://github.com/kevinjtu/BaltimoreIncinerators. Additional files or information are available upon request to the authors.

## Acknowledgements

The authors would like to thank the South Baltimore Community Land Trust for the Curtis Bay Continuous Emissions Monitoring data, community trail cam data, and unwavering advocacy on behalf of their constituents. In particular, thanks to the South Baltimore youth led team working on community monitoring of the trail cam data including Maynor Flores, Jose Alvarenga, Vilma Gutierrez, and Ryan Johnson. Thanks to Prof. Russell R. Dickerson and Prof. Timothy P. Canty for their valuable feedback during the course of the work.

## References

1. Sharma R, Sharma M, Sharma R, Sharma V. The impact of incinerators on human health and environment. Rev Environ Health [Internet]. 2013 Jan 1 [cited 2025 Jun 10];28(1). Available from: https://www.degruyter.com/document/doi/10.1515/reveh-2012-0035/html

2. Xing YF, Xu YH, Shi MH, Lian YX. The impact of PM2.5 on the human respiratory system. J Thorac Dis. 2016 Jan;8(1):E69–74.

3. Hakeem KR, Sabir M, Ozturk M, Akhtar MS, Ibrahim FH. Nitrate and Nitrogen Oxides: Sources, Health Effects and Their Remediation. Rev Environ Contam Toxicol. 2017;242:183–217.

4. Brender JD, Maantay JA, Chakraborty J. Residential Proximity to Environmental Hazards and Adverse Health Outcomes. Am J Public Health. 2011 Dec;101(S1):S37–52.

5. Zhang K. An Alternative Approach to Post-industrial Rejuvenation in Westport Waterfront, Baltimore. Azmi R, Kumar S, Akhtar N, editors. SHS Web Conf. 2022;148:03034.

6. George D. Thurston. Regarding The Public Health Impacts Of Air Emissions From The Wheelabrator Facility [Internet]. Chesapeake Bay Foundation; 2017 Nov. Available from: https://www.cbf.org/document-library/cbf-reports/thurston-wheelabrator-health-impacts-2017.pdf

7. Aubourg MA, Sawtell G, Deanes L, Fabricant N, Thomas M, Spicer K, et al. Community-driven research and capacity building to address environmental justice concerns with industrial air pollution in Curtis Bay, South Baltimore. Front Epidemiol. 2023 Sep 12;3:1198321.

8. State of Maryland, Office of the Attorney General, Environmental Crimes Unit. Plea Agreement Letter re: State of Maryland v. Curtis Bay Energy, LP [Internet]. Baltimore, MD: Office of the Attorney General, State of Maryland; 2023 May. Available from: https://www.marylandattorneygeneral.gov/news%20documents/101723_CBE.pdf

9. U.S. Environmental Protection Agency. Complaint Under Title VI of the Civil Rights Act of 1964, 42 U.S.C. § 2000d, and 40 C.F.R. Part 7 against the City of Baltimore and its agency the Baltimore City Department of Public Works, Case No. 03RNO-24-R3. 2024.

10. City of Baltimore. 10-Year Solid Waste Management Plan [Internet]. 2023 Feb. Available from: https://publicworks.baltimorecity.gov/sites/default/files/Baltimore%20SWMP%20Update_90%20DRAFT.pdf

11. U.S. Environmental Protection Agency. AP-42 Section 2.3. In: Medical Waste Incineration. 5th ed. 1993.

12. U.S. Environmental Protection Agency. Draft Technical Support Document: Maryland Area Designations for the 2010 SO2 Primary National Ambient Air Quality Standard [Internet]. U.S. Environmental Protection Agency; 2016. Available from: https://www.epa.gov/sites/default/files/2016-03/documents/md-epa-tsd-r2.pdf

13. Bio-Medical Incinerator (Capacity: 100 kg/hour or more) - Technical Specifications. [Internet]. Available from: https://www.epa.gov/sites/default/files/2016-03/documents/md-epa-tsd-r2.pdf

14. Wejkowski R, Kalisz S, Tymoszuk M, Ciukaj S, Maj I. Full-Scale Investigation of Dry Sorbent Injection for NO x Emission Control and Mercury Retention. Energies. 2021 Nov;14(22):1–13.

15. Cimorelli AJ, Perry SG, Venkatram A, Weil JC, Paine RJ, Wilson RB, et al. AERMOD: A Dispersion Model for Industrial Source Applications. Part I: General Model Formulation and Boundary Layer Characterization. J Appl Meteorol. 2005 May 1;44(5):682–93.

16. Chen J, Hoek G. Long-term exposure to PM and all-cause and cause-specific mortality: A systematic review and meta-analysis. Environ Int. 2020 Oct;143:105974.

17. Mustafić H, Jabre P, Caussin C, Murad MH, Escolano S, Tafflet M, et al. Main Air Pollutants and Myocardial Infarction: A Systematic Review and Meta-analysis. JAMA. 2012 Feb 15;307(7):713.

18. Alexeeff SE, Liao NS, Liu X, Van Den Eeden SK, Sidney S. Long-Term PM_2.5_ Exposure and Risks of Ischemic Heart Disease and Stroke Events: Review and Meta-Analysis. J Am Heart Assoc. 2021 Jan 5;10(1):e016890.

19. Ni R, Su H, Burnett RT, Guo Y, Cheng Y. Long-term exposure to PM2.5 has significant adverse effects on childhood and adult asthma: A global meta-analysis and health impact assessment. One Earth. 2024 Nov;7(11):1953–69.

20. Huang F, Pan B, Wu J, Chen E, Chen L. Relationship between exposure to PM2.5 and lung cancer incidence and mortality: A meta-analysis. Oncotarget. 2017 Jun 27;8(26):43322–31.

21. He D, Wu S, Zhao H, Qiu H, Fu Y, Li X, et al. Association between particulate matter 2.5 and diabetes mellitus: A meta-analysis of cohort studies. J Diabetes Investig. 2017 Sep;8(5):687–96.

22. Park J, Kim HJ, Lee CH, Lee CH, Lee HW. Impact of long-term exposure to ambient air pollution on the incidence of chronic obstructive pulmonary disease: A systematic review and meta-analysis. Environ Res. 2021 Mar;194:110703.

23. Huangfu P, Atkinson R. Long-term exposure to NO2 and O3 and all-cause and respiratory mortality: A systematic review and meta-analysis. Environ Int. 2020 Nov;144:105998.

24. Lee S, Tian D, He R, Cragg JJ, Carlsten C, Giang A, et al. Ambient air pollution exposure and adult asthma incidence: a systematic review and meta-analysis. Lancet Planet Health. 2024 Dec;8(12):e1065–78.

25. Hamra GB, Laden F, Cohen AJ, Raaschou-Nielsen O, Brauer M, Loomis D. Lung Cancer and Exposure to Nitrogen Dioxide and Traffic: A Systematic Review and Meta-Analysis. Environ Health Perspect. 2015 Nov;123(11):1107–12.

26. Orellano P, Reynoso J, Quaranta N. Short-term exposure to sulphur dioxide (SO2) and all-cause and respiratory mortality: A systematic review and meta-analysis. Environ Int. 2021 May;150:106434.

27. Chen K, Breitner S, Wolf K, Stafoggia M, Sera F, Vicedo-Cabrera AM, et al. Ambient carbon monoxide and daily mortality: a global time-series study in 337 cities. Lancet Planet Health. 2021 Apr;5(4):e191–9.

28. Sacks JD, Lloyd JM, Zhu Y, Anderton J, Jang CJ, Hubbell B, et al. The Environmental Benefits Mapping and Analysis Program - Community Edition (BenMAP-CE): A tool to estimate the health and economic benefits of reducing air pollution. Environ Model Softw Environ Data News. 2018 Feb 11;104:118–29.

29. United States Environmental Protection Agency. Mortality Risk Valuation [Internet]. [cited 2025 May 27]. Available from: https://www.epa.gov/environmental-economics/mortality-risk-valuation

30. Flanagan BE, Gregory EW, Hallisey EJ, Heitgerd JL, Lewis B. A Social Vulnerability Index for Disaster Management. J Homel Secur Emerg Manag [Internet]. 2011 Jan 5 [cited 2025 Jun 10];8(1). Available from: https://www.degruyter.com/document/doi/10.2202/1547-7355.1792/html

31. Maryland Department of the Environment [Internet]. 2024. Maryland Department of the Environment, Attorney General File Complaint in Circuit Court Against Curtis Bay Energy Medical Waste Incinerator for Air Pollution Violations. Available from: https://mde.maryland.gov/Documents/Complaint_filed_3-14-2024.pdf

32. Lelieveld J, Evans JS, Fnais M, Giannadaki D, Pozzer A. The contribution of outdoor air pollution sources to premature mortality on a global scale. Nature. 2015 Sep;525(7569):367–71.

33. Brown LT. The black butterfly: the harmful politics of race and space in America. Baltimore: Johns Hopkins University Press; 2021. 374 p.

34. Zhang J (Jim), Wei Y, Fang Z. Ozone Pollution: A Major Health Hazard Worldwide. Front Immunol. 2019 Oct 31;10:2518.

35. Zhou X, Zhou X, Wang C, Zhou H. Environmental and human health impacts of volatile organic compounds: A perspective review. Chemosphere. 2023 Feb;313:137489.

36. Tait PW, Brew J, Che A, Costanzo A, Danyluk A, Davis M, et al. The health impacts of waste incineration: a systematic review. Aust N Z J Public Health. 2020 Feb;44(1):40–8.

37. East Yard Communities for Env’t. Justice v. U.S. Env’t. Prot. Agency. 2023.

38. Prüss A, Emmanuel J, Stringer R, Pieper U, Townend W, Wilburn S, et al. Safe management of wastes from health-care activities / edited by A. Prüss…[et al] [Internet]. 2nd ed. Geneva: World Health Organization; 2014 [cited 2025 May 27]. Available from: https://iris.who.int/handle/10665/85349

39. Clean Air Baltimore. Medical Waste Incineration is Obsolete and Unneeded [Internet]. Available from: https://www.cleanairbmore.org/uploads/MedicalWasteIncineration.pdf

